# Psychometric Properties and Prognostic Value of the UCSD Shortness of Breath Questionnaire in Hypersensitivity Pneumonitis: A Prospective Cohort Study

**DOI:** 10.1101/2025.09.24.25336538

**Authors:** Jeffrey J. Swigris, Teng Moua, Sachin Chaudhary, Tracy N. Adams, Ayodeji Adegunsoye, Mary Beth Scholand, Namita Sood, Brian Vestal, Evans R. Fernández Pérez

## Abstract

**Rationale:** Dyspnea is a prominent symptom of hypersensitivity pneumonitis (HP), limiting patients’ activity and impairing their quality of life. The University of California San Diego Shortness of Breath questionnaire (UCSD) is a 24-item instrument used to assess dyspnea severity in patients with various respiratory conditions.

**Objective:** To examine the psychometric properties of the UCSD and assess the validity of its scores as measures of dyspnea severity and prognostic value in a prospective cohort with HP.

**Methods:** We evaluated the reliability, validity, and responsiveness of UCSD scores and assessed the association between score change and survival in a cohort of patients with HP who completed the UCSD and other HP severity metrics at baseline, 6 and 12 months. We introduce the reliable change index (RCI) and the likely change index (LCI) as ways to assess the statistical significance of within-individual change in UCSD scores and conducted analyses to provide context.

**Results:** At baseline, internal consistency (Cronbach’s coefficient alpha) was 0.97; there were significant, moderately strong correlations between UCSD scores and percent predicted forced vital capacity (FVC%) r = -0.39, percent predicted diffusing capacity of the lung for carbon monoxide (DLCO%) r = -0.33), and Borg dyspnea scores (0.55). UCSD scores were significantly different between the lowest and highest FVC%, 64.9 ± 18.9 vs. 36.2 ± 22.9. A 10- point worsening in the UCSD score was associated with a nearly 15-fold increase in time-to-death over the follow-up period.

**Conclusion:** The UCSD has acceptable psychometric properties for assessing dyspnea severity in patients with HP. Worsening dyspnea is associated with shortened survival.

## INTRODUCTION

Hypersensitivity pneumonitis (HP) is an interstitial lung disease (ILD) resulting from repeated inhalation of typically organic antigens, which drive a complex immunological reaction resulting in inflammation and often fibrosis within the small airways and lung parenchyma. Like other forms of ILD, the hallmark symptoms of HP include fatigue, cough, and activity-limiting dyspnea, any or all of which contribute to impairing their quality of life.(1, 2)

The University of California San Diego Shortness of Breath Questionnaire (UCSD) is a patient-reported outcome measure (PROM) that assesses dyspnea severity and some of its impacts through 24 items.(3) Although the UCSD was initially developed as a tool to assess dyspnea before and after completion of pulmonary rehabilitation—predominantly in patients with chronic obstructive pulmonary disease—it has been used in ILD research. In patients with idiopathic pulmonary fibrosis (IPF) and in one study that included patients with various forms of ILD (involving N=71 or 9% with HP), the UCSD demonstrated reasonably acceptable psychometric properties as a measure of dyspnea severity.(4, 5) However, there is limited evidence supporting UCSD’s prognostic power and responsiveness at baseline and over time among well-characterized patients with and without fibrotic HP, where the degree of responsiveness might differ.

We undertook this study to evaluate the reliability, validity, responsiveness, and prognostic significance of the UCSD within a prospective cohort of patients, with and without fibrotic HP.

## METHODS

### Study Population

The study cohort consisted of three independent patient populations with HP enrolled in the PREDICT-HP1 (NCT04844359) study, the PREDICT-HP2 study and the HP pirfenidone study (NCT02958917).

The PREDICT-HP1 was a multicenter, prospective observational study conducted across seven centers in the U.S. involving 137 adult participants diagnosed with fibrotic HP. The study was approved by the Mayo Clinic Central Institutional Review Board (#20-004479) and the protocol was approved by the local Institutional Review Boards prior to enrollment at each site.

The PREDICT-HP2 was an institutional review board-approved (HS#3214) prospective observational study conducted at National Jewish Health involving 52 adult participants diagnosed with non-fibrotic HP or fibrotic HP.

The HP pirfenidone study was a single-center, prospective, randomized, double-blind, placebo-controlled trial conducted at National Jewish Health involving 40 adult participants with fibrotic HP.

Participants in these three studies were aged 18 to 80 years and were required to have a documented protocol-approved multidisciplinary diagnosis of HP according to diagnostic guidelines for inclusion. They received standard care for HP, as determined by their treating physicians. The patients underwent comprehensive evaluations that included clinical assessments, responses from the UCSD, pulmonary function tests and 6-minute walk test, which included assessment of dyspnea using the Borg dyspnea index at the completion of the walk at the time of study enrollment, at 6 months and 12 months. All participants provided written informed consent.

The assessment of PROM using the UCSD in these three studies was defined as an endpoint in the protocol before patient enrollment and data collection.

### The UCSD (University of California San Diego Shortness of Breath Questionnaire)

The UCSD is a 24-item questionnaire that assesses dyspnea severity while performing each of 21 activities, and it includes another three items that ask about limitations induced by shortness of breath.(3) Each item is scored 0-5; thus, scores range from 0 to 120, with higher scores indicating greater severity of dyspnea. There is no stated recall period.

### Statistical analyses

Baseline data were summarized using counts, percentages and measures of central tendency. Analyses were hypothesis-driven (included in the Supplementary material) in accordance with COSMIN (COnsensus-based Standards for the selection of health Measurement Instruments) recommendations for studies on the measurement properties of patient-reported outcome measures.(6, 7)

Candidate anchors—variables hypothesized to be associated with dyspnea severity— included FVC (percent predicted or FVC%, the raw value in liters, and relative change in the raw value from baseline), DLCO (percent predicted or DLCO%), and the Borg dyspnea index. Analyses included the following: 1) internal consistency and test-retest reliability, 2) convergent and known-groups analyses to assess content validity, and 3) responsiveness. We also assessed the trajectory of UCSD scores over time and the association between UCSD scores and survival. Analyses were conducted in SAS, version 9.4 (SAS Institute Inc.; Cary, NC).

#### Internal consistency

Cronbach’s raw coefficient alpha was the measure of internal consistency (IC) with values > 0.7 considered acceptable.

#### Convergent and known-groups validity

We used Spearman correlations between UCSD scores and anchors at baseline to assess convergent validity. For the known-groups-at-baseline analysis, we used analysis of variance (ANOVA) with secondary, p-value corrected (Tukey) pairwise comparisons to assess the statistical significance of differences in UCSD scores between the most and least severe anchor subgroups strata. Anchors were trichotomized either to establish equal numbers within strata or to examine clinically relevant cut-points (e.g., FVC: ≤55, 55<FVC<70, or ≥70).

#### Test-retest reliability

For test-retest reliability, we used the intraclass correlation coefficient (ICC (2,1)), generated via a two-way mixed effects model for absolute agreement between baseline and 6-month UCSD scores among subjects considered stable according to FVC (FVC change in liters relative to baseline < 3%). Values > 0.7 are considered acceptable.

#### Reliable (RCI) and likely change index (LCI)

The RCI is a test of significance for change scores within an individual. The LCI is equivalent to the RCI but uses more relaxed critical values (e.g., 0.994 for p=0.32 or 0.841 for p=0.40) to allow for less than 95% confidence that change is not due to measurement error. For certain analyses, we considered changes in FVC% > 3 clinically meaningful(8, 9) and calculated the numbers of patients whose UCSD change scores met or exceeded the RCI and various LCI thresholds. We used logistic regression to generate odds ratios for meeting/exceeding the thresholds with the yes/no outcome decline in FVC% > 3.

#### Responsiveness

We used pairwise correlation, known change groups analysis, and empirical cumulative distribution function (eCDF) plots to assess the responsiveness of UCSD scores among subjects stratified by anchor change. For the correlations, we assessed pairwise Spearman coefficients between UCSD change scores and anchor change (from baseline to six months and baseline to 12 months). For the eCDF plots, we graphed the cumulative distribution of UCSD change scores from baseline to six months for tertiles of anchor change from baseline to 6 or 12 months.

#### Association between UCSD scores and time-to-death

To account for informative missingness, we analyzed the association between UCSD scores and time-to-death using a joint model that included a longitudinal submodel for UCSD scores and a survival model. Age, sex, and sex*time interaction were included as covariates in the longitudinal model that also included random effects for intercept and slope. Age, sex, FVC%, and smoking were included as covariates in the survival model that assumed a Weibull distribution.

Patients who did not die were censored at their last follow-up visit.

## RESULTS

The baseline demographic and clinical characteristics of the cohort are shown in **Table 1**. A total of 198 patients had non-missing UCSD response data at baseline, along with FVC measurements collected within 45 days of UCSD administration.

**Table 1.** Baseline demographic and clinical characteristics.

|  |  |
| --- | --- |
|  | N = 198 patients |
| Age, yrs | 68.2±8.6 |
| Female/Male | 115 (58) / 83 (42) |
| Race |  |
| Asian | 2 (1) |
| Black | 1 (<1) |
| White | 193 (97) |
| Other | 1 (<1) |
| Missing | 1 (<1) |
| FVC, L | 2.5±0.9 |
| FVC% | 71.9±19.1 |
| DLCO% | 63.1±21.3 |
| Smoker |  |
| Never | 110 (56) |
| Current or former | 88 (44) |
| UCSD score | 44.8±25.0 |
| UCSD21 score | 38.9±21.9 |
| UCSD21 category of dyspnea severity |  |
| None | 5 (2.5) |
| Very mild | 22 (11.1) |
| Mild | 62 (31.3) |
| Moderate | 42 (21.2) |
| Severe | 61 (30.8) |
| Very severe | 6 (3.0) |

Data are mean ± standard deviation or count (%) for the 198 patients with baseline UCSD absent any missing responses and FVC within 45 days of UCSD completion; FVC=forced vital capacity; DLCO=diffusing capacity of the lung for carbon monoxide; UCSD=University of California San Diego Shortness of Breath Questionnaire; UCSD21=the 21 activity items from the UCSD.

### Internal consistency

Cronbach’s alpha for baseline UCSD scores was 0.97.

### Convergent and known-groups validity

Correlations between UCSD scores and candidate anchors were moderately strong, statistically significant, and in the hypothesized direction (**Table S1** in the Supplementary Material). ANOVA confirmed statistically significant associations between UCSD scores and the three candidate anchors at baseline, and for each candidate anchor, statistically significant, p value corrected differences between least- and most-impaired strata (**Table 2**).

**Table 2.** Known groups validity analysis showing baseline UCSD and UCSD21 scores for strata of physiological variables at baseline.

| **Variable**<br>**Stratum, N** | **UCSD score** | **<u>p values</u>**<br>**Anova (top)**<br>**-- Pairwise**<br>**comparisons\*** | **UCSD21**<br>**score** | **<u>p values</u>**<br>**Anova (top)**<br>**-- Pairwise**<br>**comparisons\*** |
| --- | --- | --- | --- | --- |
| FVC, L<br>1) $\leq 1.97$, N=68<br>2) $1.97 < \text{FVC} \leq 2.69$,<br>N=65<br>3) $> 2.69$, N=65 | 55.97±25.19<br>43.26±23.55<br>34.50±21.53 | p<0.0001<br>-- 1 vs 2, 1 vs 3 * | 49.01±21.59<br>37.21±20.64<br>29.87±18.98 | p<0.0001<br>-- 1 vs 2, 1 vs 3 * |
| FVC%<br>1) $\leq 55$, N=35<br>2) $55 < \text{FVC}\% \leq 70$,<br>N=63<br>3) $> 70$, N=100 | 64.94±18.95<br>46.98±24.40<br>36.28±22.98 | p<0.0001<br>-- all pairwise<br>comparisons * | 55.94±15.99<br>40.84±21.55<br>31.63±20.30 | p<0.0001<br>-- all pairwise<br>comparisons * |
| DLCO%<br>1) $\leq 55$, N=76<br>2) $55 < \text{DLCO}\% \leq 70$,<br>N=53<br>3) $> 70$, N=61 | 51.21±25.33<br>41.71±22.33<br>37.08±24.43 | p=0.028<br>-- 1 vs 3 * | 44.47±22.25<br>36.13±19.44<br>32.18±21.35 | p=0.0029<br>-- 1 vs 3 * |
| Borg dyspnea<br>1) $\leq 2.5$, N=47<br>2) $2.5 < \text{Borg} \leq 4$, N=62<br>3) $> 4$, N=31 | 26.25±19.08<br>51.30±22.97<br>59.09±27.64 | p<0.0001<br>-- 1 vs 2, 1 vs 3 * | 22.85±16.92<br>44.33±19.79<br>51.41±24.29 | p<0.0001<br>-- 1 vs 2, 1 vs 3 * |

\*Statistically significant adjusted for multiplicity using Tukey's Honestly Significant Difference; FVC=forced vital capacity; DLCO=diffusing capacity of the lung for carbon monoxide; Note: FVC in tertiles, FVC% and DLCO% in clinically meaningful strata.

**Table 3.** Number and proportion of patients within FVC% change strata whose UCSD change score exceeded RCI or LCI thresholds.

|  | At 6 months, proportion (%) that exceeded UCSD change thresholds for significant worsening |  |  |  | At 12 months, proportion (%) that exceeded UCSD change thresholds for significant worsening |  |  |  |
| --- | --- | --- | --- | --- | --- | --- | --- | --- |
| | RCI Worse $\geq 12$ points | LCI (90%) Worse $\geq 10$ points | LCI (68%) Worse $\geq 6$ points | LCI (60%) Worse $\geq 5$ points | RCI Worse $\geq 12$ points | LCI (90%) Worse $\geq 10$ points | LCI (68%) Worse $\geq 6$ points | LCI (60%) Worse $\geq 5$ points |
| FVC% improved by 3 or more | 1/34<br>(2.9) | 2/34<br>(5.9) | 4/34<br>(11.8) | 5/34<br>(14.7) | 2/35<br>(5.7) | 3/35<br>(8.6) | 3/35<br>(8.6) | 5/35<br>(14.3) |
| $-3 < \Delta\text{FVC}\% \leq 3$ | 5/49<br>(10.2) | 5/49<br>(10.2) | 10/49<br>(20.4) | 12/49<br>(24.5) | 6/34<br>(17.7) | 7/34<br>(20.6) | 9/34<br>(26.5) | 9/34<br>(26.5) |
| FVC% worsened by $> 3$ | 11/40<br>(27.5) | 12/40<br>(30.0) | 18/40<br>(45.0) | 19/40<br>(47.5) | 9/29<br>(31.0) | 9/29<br>(31.0) | 15/29<br>(51.7) | 15/29<br>(51.7) |
| The number (%) whose FVC% worsened by $> 3$ among those whose UCSD score worsened by the threshold | 11/17<br>(64.7) | 12/19<br>(63.1) | 18/32<br>(56.3) | 19/36<br>(52.8) | 9/17<br>(52.9) | 9/19<br>(47.3) | 15/27<br>(55.6) | 15/29<br>(51.7) |
| Odds Ratio (95% CI) for FVC% worsened by $> 3$ (vs. not) | 4.8<br>(1.6-14.3) | 4.6<br>(1.6-13.0) | 4.0<br>(1.7-9.4) | 3.5<br>(1.5-7.9) | 3.4<br>(1.1-10.0) | 2.6<br>(0.9-7.4) | 5.0<br>(1.9-13.2) | 4.2<br>(1.6-10.7) |

**Table 4.** Six-month responsiveness analysis showing UCSD and UCSD21 change scores for tertiles of change scores for physiological variables.

| **<u>Variable</u>**<br>**Stratum, N** | **UCSD score** | **<u>p values</u>**<br>**Anova (top)**<br>**-- Pairwise**<br>**comparisons\*** | **UCSD21**<br>**score** | **<u>p values</u>**<br>**Anova (top)**<br>**-- Pairwise**<br>**comparisons\*** |
| --- | --- | --- | --- | --- |
| ΔFVC, L<br>1) $\leq -0.05$, N=41<br>2) $-0.05 < \Delta$FVC $\leq 0.1$,<br>N=41<br>3) $> 0.1$, N=41 | 4.85±10.77<br>0.39±13.01<br>-4.31±17.69 | P=0.01<br>-- 1 vs 3 * | 3.39±9.11<br>0.73±11.40<br>-3.19±15.19 | P=0.05 |
| ΔFVC%<br>1) $\leq -2$, N=43<br>2) $-2 < \Delta$FVC% $\leq 3$,<br>N=40<br>3) $> 3$, N=40 | 6.69±12.26<br>-2.05±10.10<br>-4.20±17.95 | P=0.001<br>-- 1 vs 2, 1 vs 3 * | 5.20±10.30<br>-1.65±9.32<br>-3.00±15.35 | P=0.004<br>-- 1 vs 2, 1 vs 3 * |
| ΔFVC (relative to baseline)<br>1) $\leq -2$, N=42<br>2) $-2 < \Delta$FVC $\leq 5$, N=42<br>3) $> 5$, N=39 | 5.35±11.08<br>-0.30±11.85<br>-4.46±18.41 | P=0.008<br>-- 1 vs 3 * | 3.78±9.29<br>0.11±10.57<br>-3.23±15.84 | P=0.02<br>-- 1 vs 3 * |
| ΔDLCO%<br>1) $\leq -3$, N=44<br>2) $-3 < \Delta$DLCO $\leq 5.3$,<br>N=38<br>3) $> 70$, N=39 | 4.46±12.78<br>-3.10±14.03<br>0.56±14.69 | P=0.05 | 3.60±11.06<br>-2.42±12.02<br>0.48±12.66 | P=0.08 |
| ΔBorg dyspnea<br>1) $\leq -1$, N=27<br>2) $-1 < \Delta$Borg $\leq 0.5$,<br>N=25<br>3) $> 0.5$, N=22 | -5.51±12.40<br>0.12±11.07<br>4.86±17.40 | P=0.03<br>-- 1 vs 3 * | -4.66±10.77<br>0.64±10.49<br>4.59±14.21 | P=0.03<br>-- 1 vs 3 * |

\*Statistically significant adjusted for multiplicity using Tukey's Honestly Significant Difference; FVC=forced vital capacity; DLCO=diffusing capacity of the lung for carbon monoxide; Note: Change is from baseline to six months; mathematically: baseline minus six months, except relative FVC change which is ((baseline minus six months)/baseline) x 100.

### Test-retest reliability

The test-retest reliability coefficient at six months was 0.84 (95% confidence interval 0.79-0.88).

### Reliable (RCI) and likely change index (LCI)

The RCIs and the LCIs for 90%, 68%, and 60% confidence intervals were 12, 10, 6, and 5, respectively. As the critical value decreased, an increasing number of patients’ change scores exceeded threshold values (Table S2 in the Supplemental Materials). At both 6 and 12 months, a decline in FVC% by more than 3 was associated with an increased odds of UCSD scores exceeding the various RCI or LCI thresholds for statistically significant change.

### Responsiveness

Correlations among change variables were weak at six months and moderately strong at 12 months (**Tables S3** and **S4**). Results for ANOVA analyses of change from baseline to six and 12 months are displayed in **Tables 4** and **S5**, respectively. At six months, the association between UCSD change and DLCO% change was not significant. At 12 months, associations between UCSD change and all three anchors were statistically significant, and for each candidate anchor, there were statistically significant p-value corrected differences between the least- and most-impaired strata. eCDF plots reveal an element of responsiveness of the UCSD by showing the proportions of patients within anchor change strata whose UCSD scores changed by various amounts at 6 (**Figure 1, Panel A**) and 12 months (**Figure 1, Panel B**). For example, greater than 60% of patients whose FVC declined by 10% or more from baseline to six months had a UCSD score that worsened by at least 6 points (LCI 68%), whereas only about 20% of patients whose FVC did not decline from baseline to six months had a UCSD score that worsened by at least 6 points.

**Figure 1.**
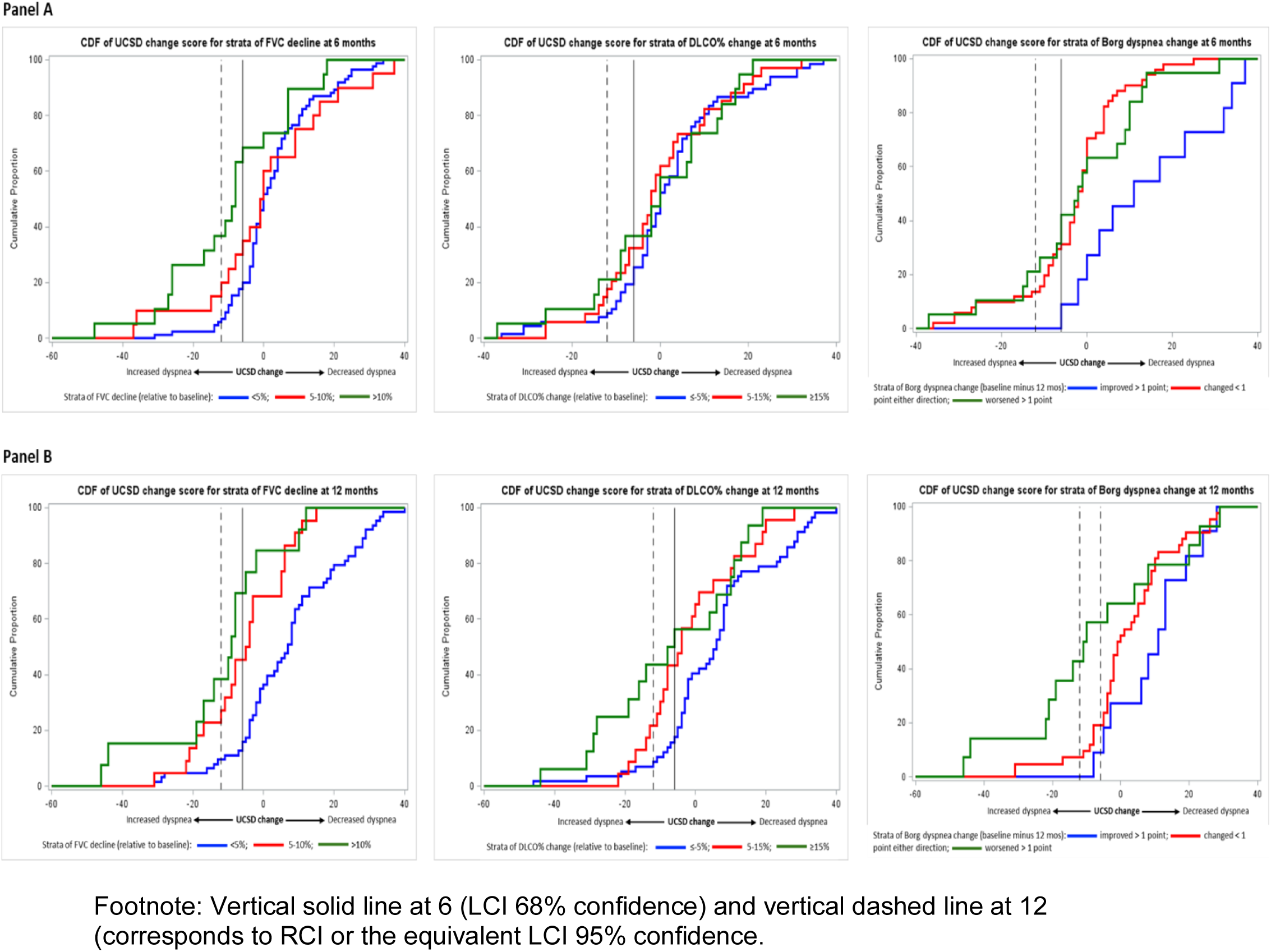
**Panel A.** eCDF plot of 6-month UCSD change scores for strata of 6-month anchor change. **Panel B.** eCDF plot of 6-month UCSD change scores for strata of 12-month anchor change.

### Association between UCSD scores and time-to-death

The longitudinal submodel of the joint model (**Table 5**) showed the following: (1) males start with lower UCSD scores than females; (2) in the cohort as a whole, modeled UCSD scores do not change significantly over 12 months; and (3) there is no difference in change over time in UCSD scores between males and females. Eighteen patients with non-missing UCSD responses at baseline and FVC within 45 days died. Median follow-up was 1.05 (IQR 0.67-1.74) years, and median survival was 4.94 (IQR 2.98-6.95) years. The survival model of the joint model revealed that FVC, smoking status, and UCSD slope were associated with survival; any 10-point increase in UCSD score over 12 months was associated with a nearly 15-fold increase in the risk of death during follow-up.

**Table 5.**
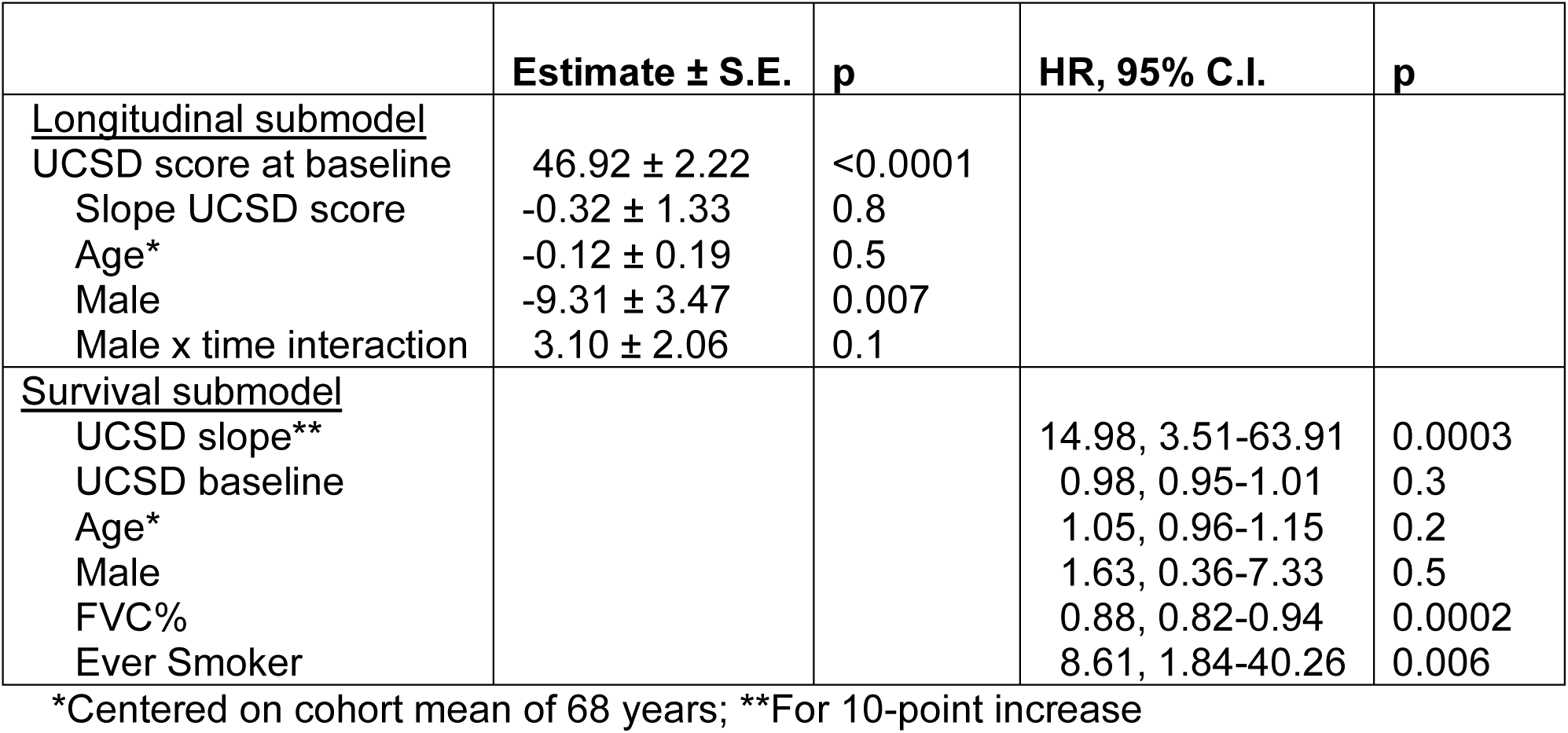
Results of joint model for UCSD score over time and survival.

## DISCUSSION

We conducted analyses to assess the psychometric properties of the UCSD as a measure of dyspnea severity in a cohort of patients with fibrotic and nonfibrotic HP and found (1) Internal consistency was high, confirming the strong relationship among its 24 items. This suggests all items are effectively measuring the same general construct: dyspnea severity, and (2) a 10-point increase in UCSD score over 12 months was associated with mortality.

The weak to moderate correlation between changes in PROM scores and changes in physiological measures—a very well-established finding across multiple studies in ILD(10, 11)—dampens confidence in results of many analyses requiring anchor values, including test-retest and responsiveness. However, the test-retest correlation coefficient at six months far exceeded the acceptability value of 0.7. In this type of study, investigators are forced to conduct test-retest analyses a bit unconventionally: Ideally, test-retest is carried out over a short period of time; two weeks is typical. By having patients complete the PROM at baseline and then two weeks later, they are likely to be clinically stable during this timeframe, and enough time has passed to minimize the risk of them recalling and simply repeating their earlier responses.

We elected not to estimate within-patient meaningful change thresholds because correlations between change variables at 6 months did not meet our *a priori* threshold (0.371), and the number of patients whose anchor values changed limited power for analyses at 12 months.

There is ongoing controversy and uncertainty in the QOL literature around how to estimate within-individual and between-groups meaningful change.(12–18) One group of experts suggests using a statistical test on change scores within an individual, arguing that a statistically significant change in a PROM score is almost certainly going to be meaningful to the individual. The reliable change index (RCI) yields the threshold for significant change (at the 95% confidence level) in a PROM score within an individual; a score that exceeds the threshold is unlikely to be due to measurement error and thus likely to represent true change (at 95% confidence).(13) Because within-individual change in PROM score may not be a high-stakes issue, investigators may wish to liberalize the threshold to increase sensitivity and capture patients who are more likely than not to have truly changed (e.g., 90%, 68%, 60%, or even 51%); this is the value provided by the LCI. Interestingly and somewhat reassuringly, the threshold values we calculated for the RCI and LCIs (12, 10, 6, and 5 points) align with published minimal important differences for the UCSD in idiopathic pulmonary fibrosis (IPF).(4, 5)

In longitudinal studies, missing data are typically unavoidable. Patient subjects may miss study visits or not complete outcome assessments for various reasons. Missing visits because the disease has progressed, and patients are too dyspneic, frail, or need too much oxygen, or because a patient has died, creates nonignorable missingness. We used a joint model to account for this nonignorable missingness in UCSD scores while assessing their association with time-to-death. We found that a 10-point increase in UCSD score (which essentially corresponds to a 90% confidence that true change has occurred) is associated with a nearly 15-fold increase in death over the follow-up period.

The study’s strengths include the prospective design, the protocolized aim of evaluating PROM with the UCSD prior to the start of the study, adjusting for the presence of lung fibrosis, and the standardized HP diagnoses and assessments conducted within a multicenter cohort, enhancing the generalizability of findings.

Our study has limitations. As noted above, 2-week response data were not collected, so a test-retest was performed at six months. Correlations between UCSD change and candidate anchor change failed to meet our *a priori* threshold, so we did not attempt to generate an estimate for important change. However, we introduce the RCI and LCI as a way to assess the likelihood of measurement error vs. true change in an individual’s scores across time points.

To illustrate this concept, consider an LCI at the 68% confidence level: this corresponds to a p-value of 0.32. Thus, under conditions of the null (i.e., true change in UCSD score has not occurred), the probability of seeing an increase of 6 or more in UCSD score is 32%. Because this is a “low” percent, we reject the null and accept the alternative – true change has occurred. The advantage of the LCI is that confidence can be set according to the preferences of the investigator. However, it is important to note that the RCI and LCI focus on statistical significance and do not directly address the issue of meaningfulness.

Despite these limitations, the results can be considered a foundation of validity for the UCSD score as a measure of dyspnea severity in patients with HP. There is no question the UCSD has face validity as a dyspnea severity measure. Our results show that internal consistency, test-retest, and construct validity appear acceptable in this target population, and change in UCSD score is associated with increased risk of death. Scores from another questionnaire, the Dyspnea domain of L-PF-35 (the Living with Pulmonary Fibrosis 35-item questionnaire) has been shown to possess validity as a measure of dyspnea severity in patients with HP.(19) Validation is a never-ending process. It is used to build confidence in inferences generated from PROM scores about patients in the target population – the question is not whether the PROM is valid but whether the inferences drawn from its score are valid. Thus, additional research is needed to further build on the foundation of validity established by this study. Also, if of interest, more studies will be required to assess whether the UCSD or L-PF better captures dyspnea severity (within-individual, between-groups, and over time) in patients with HP.

## Data Availability

All data produced in the present study are available upon reasonable request to the authors.

## Supplementary Materials

**Table S1.**
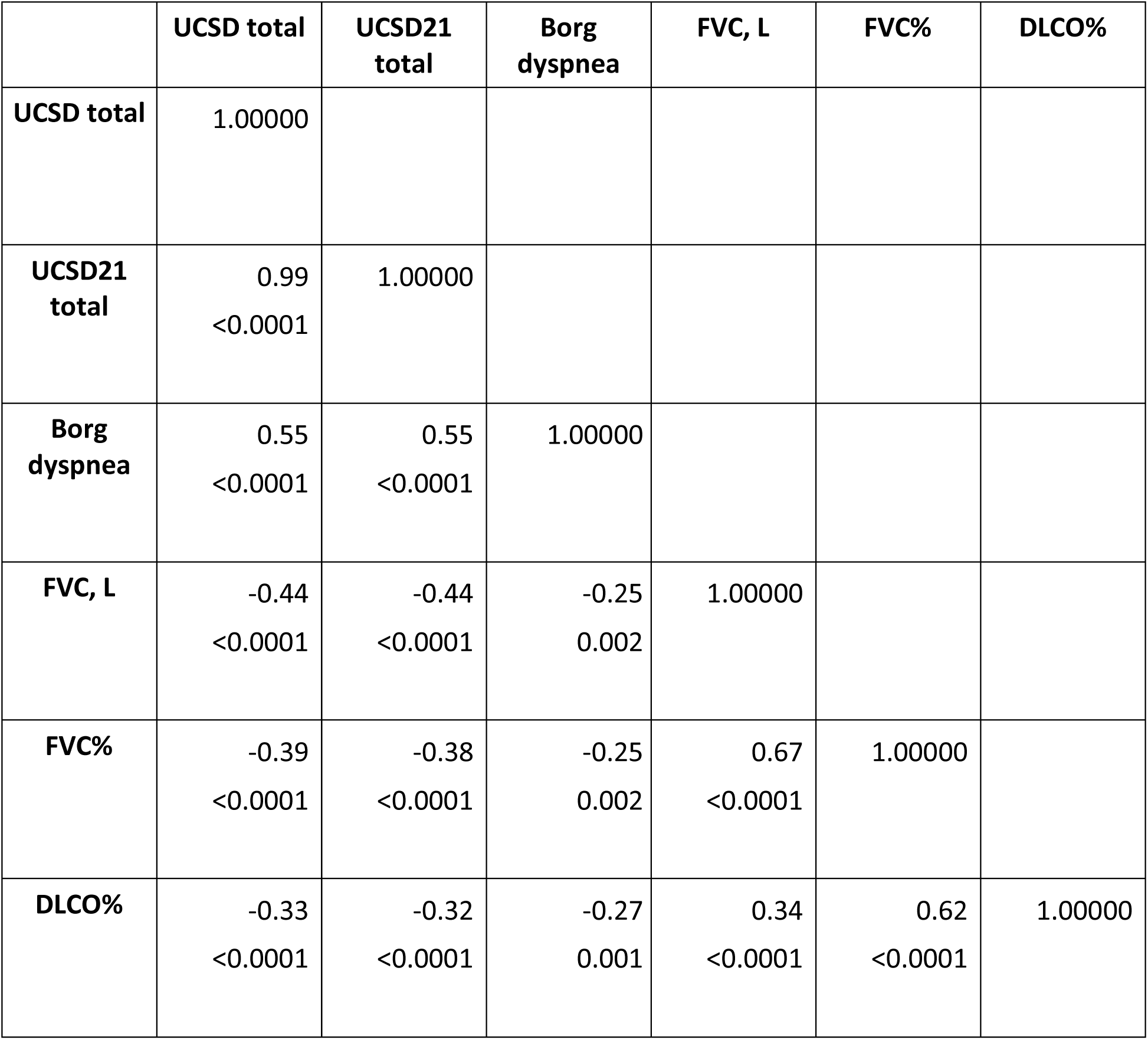
Correlation among variables at baseline.

|  | **UCSD total** | **UCSD21 total** | **Borg dyspnea** | **FVC, L** | **FVC%** | **DLCO%** |
| --- | --- | --- | --- | --- | --- | --- |
| **UCSD total** | 1.00000 |  |  |  |  |  |
| **UCSD21 total** | 0.99<br><0.0001 | 1.00000 |  |  |  |  |
| **Borg dyspnea** | 0.55<br><0.0001 | 0.55<br><0.0001 | 1.00000 |  |  |  |
| **FVC, L** | -0.44<br><0.0001 | -0.44<br><0.0001 | -0.25<br>0.002 | 1.00000 |  |  |
| **FVC%** | -0.39<br><0.0001 | -0.38<br><0.0001 | -0.25<br>0.002 | 0.67<br><0.0001 | 1.00000 |  |
| **DLCO%** | -0.33<br><0.0001 | -0.32<br><0.0001 | -0.27<br>0.001 | 0.34<br><0.0001 | 0.62<br><0.0001 | 1.00000 |

**Table S2.** Reliable (RCI) and likely change index (LCI) for worsening of UCSD score at various critical values.

| **Confidence interval** | **Critical value** | **UCSD change threshold for significant change at the corresponding critical value** | **6 months**<br>**N (%)**<br>**exceeding threshold** | **12 months**<br>**N (%)**<br>**exceeding threshold** |
| --- | --- | --- | --- | --- |
| 95% (RCI) | 1.96 | 12 | 17 (13.7) | 17 (17.3) |
| 90% (LCI) | 1.645 | 10 | 20 (16.1) | 19 (19.3) |
| 68% (LCI) | 0.994 | 6 | 33 (26.6) | 27 (27.5) |
| 60% (LCI) | 0.841 | 5 | 37 (29.8) | 29 (29.5) |

**Table S3.** Correlation among change variables at 6 months.

|  | **ΔUCSD** | **ΔUCSD21** | **ΔBorg**<br>**dyspnea** | **ΔFVC** | **ΔFVC%** | **ΔFVC**<br>**(relative**<br>**to**<br>**baseline)** | **ΔDLCO%** |
| --- | --- | --- | --- | --- | --- | --- | --- |
| **ΔUCSD** | 1.00000 |  |  |  |  |  |  |
| **ΔUCSD21** | 0.97<br><.0001 | 1.00000 |  |  |  |  |  |
| **ΔBorg**<br>**dyspnea** | 0.27<br>0.01 | 0.30<br>0.007 | 1.00000 |  |  |  |  |
| **ΔFVC** | -0.28<br>0.001 | -0.27<br>0.002 | -0.08<br>0.4 | 1.00000 |  |  |  |
| **ΔFVC%** | -0.27<br>0.001 | -0.26<br>0.003 | -0.11<br>0.3 | 0.95<br><0.0001 | 1.00000 |  |  |
| **ΔFVC**<br>**(relative**<br>**to**<br>**baseline)** | -0.26<br>0.003 | -0.24<br>0.006 | -0.05<br>0.6 | 0.97<br><0.0001 | 0.93<br><0.0001 | 1.00000 |  |
| **ΔDLCO%** | -0.18<br>0.04 | -0.15<br>0.08 | -0.04<br>0.6 | 0.39<br><0.0001 | 0.40<br><0.0001 | 0.38<br><0.0001 | 1.00000 |

| | ΔUCSD | ΔUCSD21 | ΔBorg<br>dyspnea | ΔFVC | ΔFVC% | ΔFVC<br>(relative<br>to<br>baseline) | ΔDLCO% |
| --- | --- | --- | --- | --- | --- | --- | --- |

**Table S4.** Correlation among change variables at 12 months.

| | ΔUCSD | ΔUCSD21 | ΔBorg<br>dyspnea | ΔFVC | ΔFVC% | ΔFVC<br>(relative<br>to<br>baseline) | ΔDLCO% |
| --- | --- | --- | --- | --- | --- | --- | --- |
| ΔUCSD | 1.00000 | | | | | | |
| ΔUCSD21 | 0.98<br><0.0001 | 1.00000 | | | | | |
| ΔBorg<br>dyspnea | 0.33<br>0.008 | 0.30<br>0.01 | 1.00000 | | | | |
| ΔFVC | -0.38<br><0.0001 | -0.37<br>0.0002 | -0.27<br>0.03 | 1.00000 | | | |
| ΔFVC% | -0.39<br><0.0001 | -0.37<br>0.0001 | -0.25<br>0.05 | 0.93<br><0.0001 | 1.00000 | | |
| ΔFVC<br>(relative<br>to<br>baseline) | -0.40<br><0.0001 | -0.39<br><0.0001 | -0.240<br>0.05 | 0.97<br><0.0001 | 0.93<br><0.0001 | 1.00000 | |
| ΔDLCO% | -0.35<br>0.0004 | -0.37<br>0.0003 | -0.11<br>0.3 | 0.22<br>0.02 | 0.25<br>0.01 | 0.21<br>0.03 | 1.00000 |

**Table S5.** Twelve-month responsiveness analysis showing UCSD and UCSD21 change scores for tertiles of change scores for physiological variables.

| <u>Variable</u><br>Stratum, N | UCSD score | <u>p values</u><br>Anova (top)<br>-- Pairwise<br>comparisons* | UCSD21 score | <u>p values</u><br>Anova (top)<br>-- Pairwise<br>comparisons* |
| --- | --- | --- | --- | --- |
| ΔFVC, L<br>1) $\leq -0.08$, N=33<br>2) $-0.08 < \Delta$FVC $\leq 0.14$, N=32<br>3) $> 0.1$, N=33 | 8.57±14.15<br>3.15±117.23<br>-6.63±15.38 | P=0.0006<br>-- 1 vs 3, 2 vs 3 * | 6.30±13.08<br>2.56±14.58<br>-5.57±13.49 | P=0.002<br>-- 1 vs 3, 2 vs 3 * |
| ΔFVC%<br>1) $\leq -4$, N=33<br>2) $-4 < \Delta$FVC% $\leq 2$, N=29<br>3) $> 2$, N=36 | 8.12±14.56<br>3.10±18.06<br>-5.36±15.09 | P=0.002<br>-- 1 vs 3* | 5.90±12.96<br>2.20±15.76<br>-4.25±13.27 | P=0.01<br>-- 1 vs 3 * |
| ΔFVC (relative to baseline)<br>1) $\leq -2.7$, N=34<br>2) $-2.7 < \Delta$FVC $\leq 5.2$, N=31<br>3) $> 5.2$, N=33 | 9.88±14.16<br>2.74±16.88<br>-7.75±14.44 | P<0.0001<br>-- 1 vs 3, 2 vs 3 * | 7.52±13.09<br>2.25±14.30<br>-6.67±12.62 | P=0.0002<br>-- 1 vs 3, 2 vs 3 * |
| ΔDLCO%<br>1) $\leq -4$, N=30<br>2) $-4 < \Delta$DLCO $\leq 5.5$, N=30<br>3) $> 5.5$, N=33 | 11.30±14.54<br>0.36±13.52<br>-3.69±16.78 | P=0.0005<br>-- 1 vs 2, 1 vs 3 * | 9.13±12.61<br>-0.16±12.09<br>-3.36±14.72 | P=0.001<br>-- 1 vs 2, 1 vs 3 * |
| ΔBorg dyspnea<br>1) $\leq -0.5$, N=18<br>2) $-0.5 < \Delta$Borg $\leq 0.0$, N=23<br>3) $> 0.0$, N=20 | -7.16±21.22<br>3.60±11.14<br>7.90±13.07 | P=0.01<br>-- 1 vs 3 * | -5.83±18.18<br>2.65±10.48<br>6.40±11.75 | P=0.02<br>-- 1 vs 3 * |

\*Statistically significant adjusted for multiplicity using Tukey's Honestly Significant Difference; FVC=forced vital capacity; DLCO=diffusing capacity of the lung for carbon monoxide; Note: Change is from baseline to six months; mathematically: baseline minus six months, except relative FVC change which is ((baseline minus six months)/baseline) x 100.

